# Protocol for a cluster randomised trial to evaluate a community-level complementary-food safety and hygiene and nutrition intervention in Mali: The MaaCiwara study

**DOI:** 10.1101/2021.12.15.21267512

**Authors:** Samuel I Watson, E Asamane, RJ Lilford, K Hemming, Cheick Sidibe, Ryan T. Rego, Sami Bensassi, Ayouba Diarra, Youssouf Diarra, Samba Diop, Om Prasad Gautam, Mohammad Sirajul Islam, Louise Jackson, Kate Jolly, Kassoum Kayentao, Ousmane Koita, Buba Manjang, Susan Tebbs, Nicola Gale, Paula Griffiths, Sandy Cairncross, Ousmane Toure, Semira Manaseki-Holland

## Abstract

**Background:** Diarrheal disease remains a significant cause of morbidity and mortality among the under-fives in many low- and middle-income countries. Changes to food safety practices and feeding methods around the weaning period, alongside improved nutrition, may significantly reduce the risk of disease and improve development for infants. This article describes a protocol for a cluster randomized trial to evaluate the effectiveness of a multi-faceted community-based educational intervention that aims to improve food safety and hygiene behaviours and enhance child nutrition.

**Methods:** We will conduct a mixed-methods, parallel cluster randomised controlled trial with baseline measures. 120 clusters comprising small urban and rural communities will be recruited in equal numbers and randomly allocated in a 1:1 ratio to either treatment or control arms. Participants will be mother-child dyads (27 per cluster period) with children aged 6 to 24 months. Data collection will comprise a day of observation and interviews with each participating mother-child pair and will take place at baseline and four and 15 months post-intervention. The primary analysis will estimate the effectiveness of the intervention on changes to complementary food safety and preparation behaviours, food and water contamination, and diarrhoea. Secondary outcomes include maternal autonomy, enteric infection, nutritional content of meals, and child anthropometry. A secondary structural equation analysis will be conducted to examine the causal relationships between the different outcomes.

**Conclusions:** The trial will provide evidence on the effectiveness of community-based behavioural change and educational interventions designed to reduce the burden of diarrhoeal disease in the under fives, and how effectiveness varies across different contexts.

## INTRODUCTION

### Background

Approximately 525,000 children under five die each year from diarrhoeal disease [1]. It remains the fifth leading cause of death among the under-fives worldwide [2], with Sub-Saharan Africa accounting for 40% of cases [1]. Besides mortality, diarrhoeal disease also contributes to infant malnutrition, stunting, and developmental delays [3, 4]. One particularly vulnerable group are children starting to eat semi-solid and solid foods (known as complementary-foods) [5].

While many diarrhoea causing pathogens are destroyed during high-heat cooking, the handling and storage of food can still contribute largely through food contamination [6]. The World Health Organisation (WHO) advocates for targeted interventions to improve complementary-food safety and hygiene, particularly through the ‘5 key steps’ of food safety, and Hazard Analysis Critical Control Points (HACCP) to identify specific ameliorating behaviours [7, 8]. Interventions to improve complementary-food safety, reduce geophagy, and improve nutritional intake are potentially of great importance for better diarrhoea control and hence growth and developmental outcomes [2, 9]. Interventions targeting these areas together could potentially bring synergy, thus enhancing behavioural reinforcement, that could reduce diarrhoea and improve nutritional status of children.

Smaller-scale Randomised Controlled Trials (RCTs) in West Africa and South East Asia exploring the safety of complementary-foods have shown that behaviour change interventions which promote complementary-food safety and hygiene can have a positive impact on child health [10-12]. However, advice and education has a limited impact on behaviour change unless accompanied by means to motivate and empower mothers in the community. Such empowerment includes community support and encouragement, and a change of social norms [13, 14]. Drama and traditional arts are known to captivate and involve communities, impart sensitive knowledge and set trends in behaviour [15]. Yet previous interventions targeting diet or diarrhoea have seldom reported to draw on cultural dramatic arts, community assets and social norms to motivate behaviour change. African communities have a particularly strong cultural heritage to underpin such potential impact. In this project, we will evaluate an intervention that seeks to change behaviours through personal motivation and social support and to empower mothers through improved social status. The intervention aims to enhance motivators for behaviour change of mothers and modify social norms through the use of cultural performing arts as well as community-level campaigns with community events, public pledging, competitions, peer education and support and engaging community leaders.

This project builds on past work examining behaviour change interventions for complementary-food hygiene in rural areas [12, 16, 17]. We will also incorporate urban areas in this study, which face different challenges with regards to child health and diarrhoea to rural areas, including population density, particularly in informal settlements, and differences in the role of women, families, and hierarchies [18, 19]. In order to extend the scope of the intervention, we have added a nutrition behaviour change component that fits naturally with the complementary-food safety and hygiene intervention at the time when the child is introduced to complementary feeding. The causal pathways through which our intervention components are hypothesised to improve clinical outcomes are represented in Figure 2. For example, diarrhoea is often a self-limiting condition but can lead to hospitalisation and enteropathy that affects growth. Growth can be improved through a nutrition behaviour change and reduced diarrhoea, while child development can improve through the mediation of both improved nutrition and reduced illness such as diarrhoea.

### Objectives

The aim of the study is to evaluate the implementation and effectiveness of the MaaCiwara complementary-food safety and hygiene and nutrition intervention. The evaluation comprises several objectives:

1. To evaluate the effectiveness of the intervention to promote improved drinking-water and complementary-food safety and hygiene behaviour to reduce water and food contamination, and observed diarrhoea.
2. To conduct a subgroup/secondary analysis to evaluate whether the effectiveness of the intervention on the primary outcomes varies between:
  a. urban and rural settings
  b. over time between 4- and 15-months post-intervention follow-up.
3. To explore the causal processes that affect the effectiveness, or lack thereof, of the intervention on key clinical endpoints, as mediated by outcomes, such as geophagy, and maternal knowledge and behaviour.
4. To conduct a secondary analysis to evaluate the effect of the intervention on dietary diversity and meal frequency, growth and development, and explore variations due between urban and rural settings.
5. To estimate the cost-effectiveness of the intervention as incremental cost per case of diarrhoea averted and per unit improvements in growth and development.

## METHODS

### Trial design

We will conduct a mixed-methods, parallel cluster randomised controlled trial with baseline measures. Clusters (N=120) will be small urban and rural communities, recruited in equal numbers, randomly allocated in a 1:1 ratio to either treatment or control arms stratified by urban/rural status. Regarding the start of trial timeline as the beginning of data collection, the data collection and intervention will be as follows: Baseline observations will be taken over 3 months (months 1-3). The main part of the intervention will be rolled out over 3-4 months in all clusters and intensely over 35 days in each cluster (months 9-12). A mid-line and an end-line set of observations will be carried out over 3 months each (months 14-16 and 25-27) Sampling within cluster-periods will be cross-sectional. (See Supplementary File 1 for SPIRIT checklist)

### Participants

#### Setting

Urban clusters will be recruited from Bamako city, the capital of Mali, while rural clusters will be recruited from villages in the surrounding Bamako, Sikaso and Sego regions.

#### Eligibility criteria and cluster design

##### Urban clusters

We created 60 urban clusters in Bamako city (Figure 1). The clusters are designed to include predominantly poor communities to be approximately comparable to the rural clusters. The clusters are areas of 500 to 2,000 people (approximately 200 to 500 households). These clusters are bounded by obvious environmental features including roads and rivers. The clusters are also designed to have a relatively central community area. As much space as possible between cluster areas was included to reduce the risk of contamination, either from individuals in one cluster visiting the intervention in another site, or knowledge produced by the intervention being communicated over larger areas.

**Figure 1.**
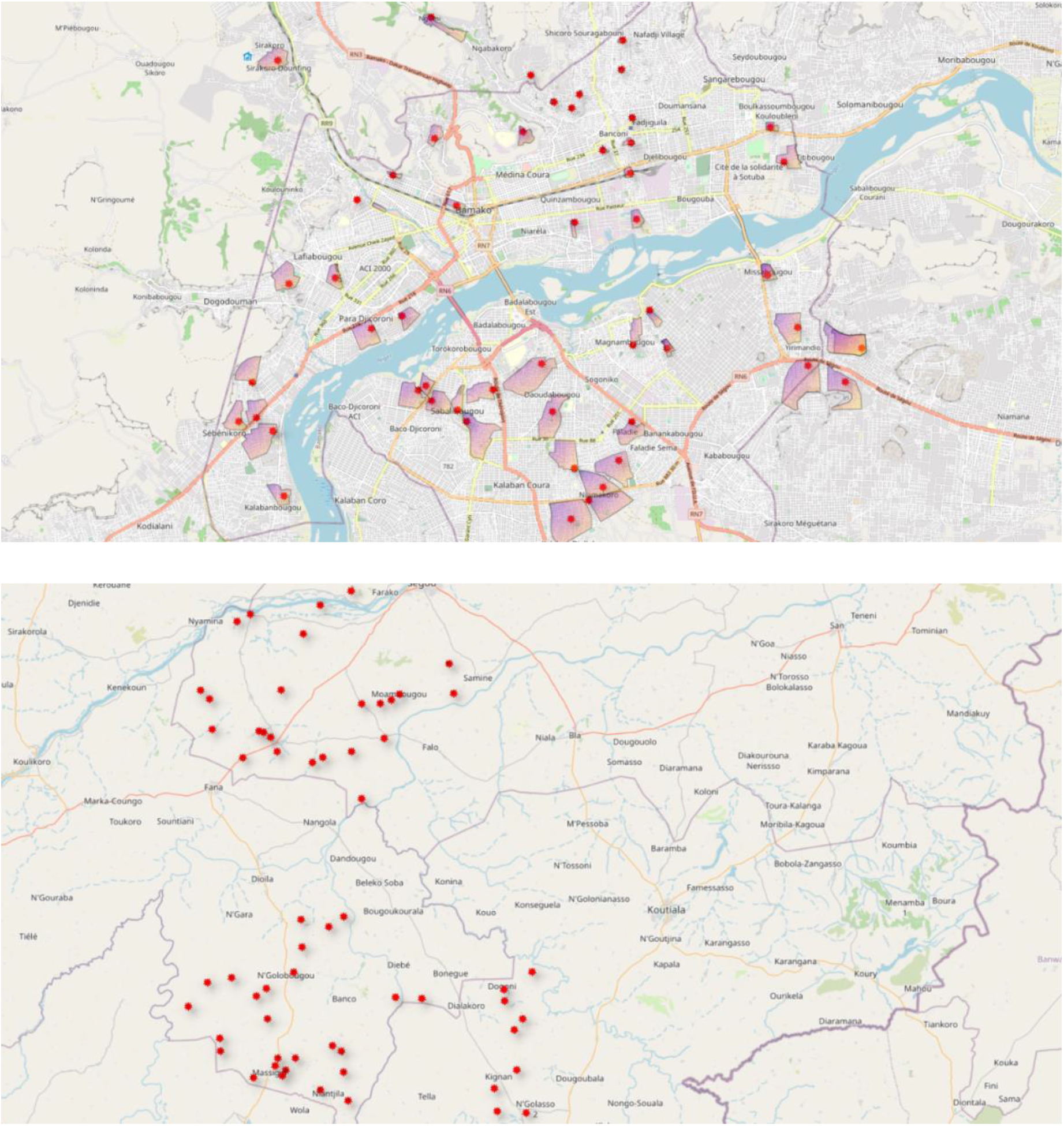
Cluster boundaries and locations in urban Bamako city (top) and rural Bamako region (bottom). Scales differ: villages in the rural region are a minimum of 5km apart.

##### Rural clusters

Sixty villages were sampled for inclusion from the population of eligible villages (population of 500 to 2,000) in Bamako region using a weighted stratified random sample. A sampling frame of all possible villages in each district was constructed. The stratification variables were village size (<=median, > median) and availability of community-led total sanitation. We determined the proportion of villages with and without each characteristic (to act as a weighting factor). Including two stratification variables each with two levels will result in four strata. The number to be sampled from each group were determined using the weighting factors. Villages were then randomly ordered within strata and approached for inclusion following this random order. Included villages are a minimum of 5km apart. The location of villages is shown in Figure 1.

##### Individual participants

Mother-child pairs will be the unit of observation in the study. Within each cluster period we will aim to recruit 27 consenting mothers with at least one child aged 6 to 24 months. (see Randomisation - Implementation)

### Intervention

The intervention will be an adapted version of a community level complementary-food hygiene and safety intervention previously carried out in Mali, Bangladesh and Nepal, and evaluated in the Gambia[17, 20]. The intervention will also include hygienic play and dietary diversity and meal frequency promotion, which were not included in latter studies, including the Gambia intervention [17, 20].

The intervention schedule and tools used in the MaaChampion Gambian study [17, 20] will be contextualised and adapted by applying the findings of a mixed-methods formative research study conducted in Mali in 2021 and developing a creative brief for a Mali creative team to design the community level intervention components. The full intervention, once adapted, will be described in a separate publication in compliance with the template for intervention description and replication (TIDieR) checklist and guide [21]. The basic components and principles of the intervention will be the same as the MaaChampion intervention.[17, 20] The adaptation will mainly influence the choice of behaviours, language and cultural aspects of dramatic arts, and the focus on community hierarchies.

Table 1 presents the provisional timeline and steps of the intervention delivery. The intervention will involve 5 days of campaign community visits dispersed across 35 days (days 1, 2, 15, 17, and 35), including home visits, community events, neighbourhood meetings, dramatic arts (songs, drama, stories, animation), and competitions based around achieving a model-mother status. Community volunteer home visits followed between each campaign day, and a reminder campaign day after nine months to embed the behaviours into social norms, and thereby enhancing sustainability. They will be paid minimally (comparable to local health system health volunteer) for the first year of the project.

**Table 1.** Example timetable of the intervention (T = days from start of intervention period)

| Visits | T | Activity or event |
| --- | --- | --- |
| Visit 1 | 1 | Meeting with the community leader |
|  |  | Community announcement to invite community members to the upcoming afternoon event |
|  |  | House-to-house visit with community mother supervisors |
|  |  | Record a short video with community leader |
|  |  | Afternoon community event (includes dancing and song, animation, street theatre, public pledging etc) |
|  |  | Training assistants of the “Mother Supervisors” |
| Visit 2 | 2 | Meeting with the community leader |
|  |  | Community announcement to invite community members to the upcoming afternoon event |
|  |  | House-to-house visit with community mother supervisors |
|  |  | Neighbourhood meeting with demonstrations and stories |
| No visit | 3-14 | “Mother Supervisors” and assistants carry out house-to-house visits to encourage and assess mothers |
| Visit 3 | 15 | Meeting with the community leader |
|  |  | Community announcement to invite community members to the upcoming afternoon event |
|  |  | House-to-house visit with community “Mother Supervisors” |
|  |  | Afternoon community event |
| No visit | 15-24 | “Mother Supervisors” and assistants carry out house-to-house visits to encourage and assess mothers |
| Visit 4 | 27 | Meeting with the community leader |
|  |  | Community announcement to invite community members to the upcoming afternoon event |
|  |  | House-to-house visit with community “Mother Supervisors” |
|  |  | Afternoon community event |
| No visit | 28-40 | “Mother Supervisors” and assistants or MaaCiwaras carry out house-to-house visits to encourage and assess mothers |
| Visit 5 | 40 | Meeting with the community leader |
|  |  | Community announcement to invite community members to the upcoming afternoon event |
|  |  | House-to-house visit with community “Mother Supervisors” |
|  |  | Afternoon community event and certification of village and mothers |
| No visit | 40-270 | “Mother Supervisors” and assistants or MaaCiwaras carry out house-to-house visits to encourage and assess mothers |
| Visit 6 (9 month) | ~9m | Meeting with the community leader |
|  |  | Community announcement to invite community members to the upcoming afternoon event |
|  |  | House-to-house visit with community “Mother Supervisors” |
|  |  | Afternoon community event |

Implementation will be through Intervention teams who will include district level community health promotion or public health staff, community traditional communicators (traditional singers, drummers, and performing artists) and respected community members as assisted volunteers (often older mothers or traditional birth attendants). All community members (including fathers) will be involved as well as community leaders in promoting the intervention.

### Control condition

The control communities will receive a 1-day community-based campaign on the use of water in homes and outdoors. The content is designed to be similar to the intervention in terms of being a community-based intervention delivered through a one day campaign event, but not contain equivalent content on food and water preparation, hygiene, child nutrition or hygienic play. It is designed to serve as a “placebo”. The specific choice of topic for control communities will be finalised after the adaptation of the intervention to ensure minimal overlap. As with the intervention, a Public Health/Health Promotion Officer will provide the visit in each control community. The intervention and control village activities will be delivered in parallel.

### Randomisation and sampling

#### Sequence generation, allocation concealment, and cluster allocation

After baseline data collection, clusters will be randomly allocated by an independent statistician in a 1:1 ratio to either treatment or control arms stratified by urban/rural status (30 intervention and 30 control clusters from both urban and rural settings). Within strata we will use a minimisation method to allocate clusters to either treatment or control arms to improve the efficiency of the design given large potential heterogeneity between the clusters. The covariates we will use are: population size (< or >= median), presence of community-led total sanitation in rural areas, and whether there is a school located in the cluster. Allocation is planned to take place immediately prior to intervention, following baseline data collection in all participating clusters, however the use of a minimisation procedure permits allocation in the event baseline data collection is running behind schedule or clusters drop out.

#### Implementation

##### Individual sampling

We will recruit 27 mother-child pairs (child aged between six and 36 months) per cluster-period. Prior to each round of data collection, local community female informants (usually volunteers working with the local health system) will be asked to compile a numbered list of all eligible mother-child pairs in the area (or approximate area in the case of urban) of the cluster. A random ordering of pairs will be generated by the trial statistician in advance of data collection and provided to the field supervisor. The field team will arrive at the cluster the day prior to planned data collection, where they will sequentially attempt to identify each mother-child pair on the list in order until a total of 27 pairs have agreed to participate.

Participating households will be assigned a predetermined unique identifier based on the cluster-period and list position. Mother-child pairs who are either not locatable, who reside outside the cluster boundary, or who are not available the following day will be excluded. In post-intervention periods it is possible some mothers may be included who were in previous rounds. Despite this, we will treat sampling a cross-sectional for three main reasons. There are logistical challenges to tracking consistent trial identifiers across trial data collection periods in this setting. Excluding mothers who have been included in prior survey rounds may significantly limit the available sample size. And statistical analyses incorporating an individual-level random effect to capture cohort effects will not likely be possible as the majority of participants will only be observed once during the study.

##### Informed Consent

Initially at the time of cluster recruitment, written and oral permission was sought from the community leader(s) within each cluster. We provided a verbal presentation of the intervention and study to the community leader(s) along with an information sheet on the study. If the community leader could not read, it was read to them by the field worker. All materials were translated into French and Bambara and presented in the language of the community leader(s). Community leaders are allowed to withdraw their communities from the study at any time.

Subject to the permission of community leaders, we will approach eligible mothers to participate in the study at each time point. The day prior to any observation or data collection prospective study participants will be approached and provided with information about the data collection in French or Bambara as required in writing or to be read/explained to them, and consent will be sought (see Supplementary File 2). Mothers are allowed to withdraw from the study at any time.

### Blinding

Due to the nature of the intervention, blinding of mothers and the intervention team will not be possible. However, the mothers will not be informed individually that there is a trial taking place since the consent was taken from community leaders in Spring of 2021. During the assessment rounds, the data collection will not be linked to the intervention or the trial. The mothers and data collectors will be masked to the nature of the assessment: At baseline, 4- and 15-month assessments, mothers will be informed that the assessment is investigating how children aged 6 to 36 months and their mothers spend their days in rural and urban Mali and to support delivery of local health and social services. Complementary-food safety and hygiene components of the assessment tools will be concealed in a larger assessment, with a package of observation tools and questionnaires about household food and water use, mother and child activities (including observation and a questionnaire on child care and play activities), health-seeking, water and sanitation, income in the household, and village activities, including sub-study on the use of plastics. Thus, the data collectors will be trained for, and mothers consented to, the conduct of this larger assessment of the household’s food and water consumption, health, and childcare.

New independent field teams will be recruited in each round and will not be informed of the intervention or the inter-village comparison. Data analysts will be also blinded as to treatment allocation.

### Outcomes

The intervention is complex in nature and is designed to target multiple causal pathways that could impact on several child health outcomes. Figure 2 shows a causal diagram indicating the assumed relationships between the different types of outcome. For our main analysis we focus on a “primary causal pathway” for complementary-food safety and hygiene linking the intervention to diarrhoeal disease via water and complementary-food safety and hygiene behaviour and contamination. From this pathway we include three primary outcomes, one behavioural, one microbiological, and one clinical (to capture behaviour -> exposure -> incident case).

**Figure 2.**
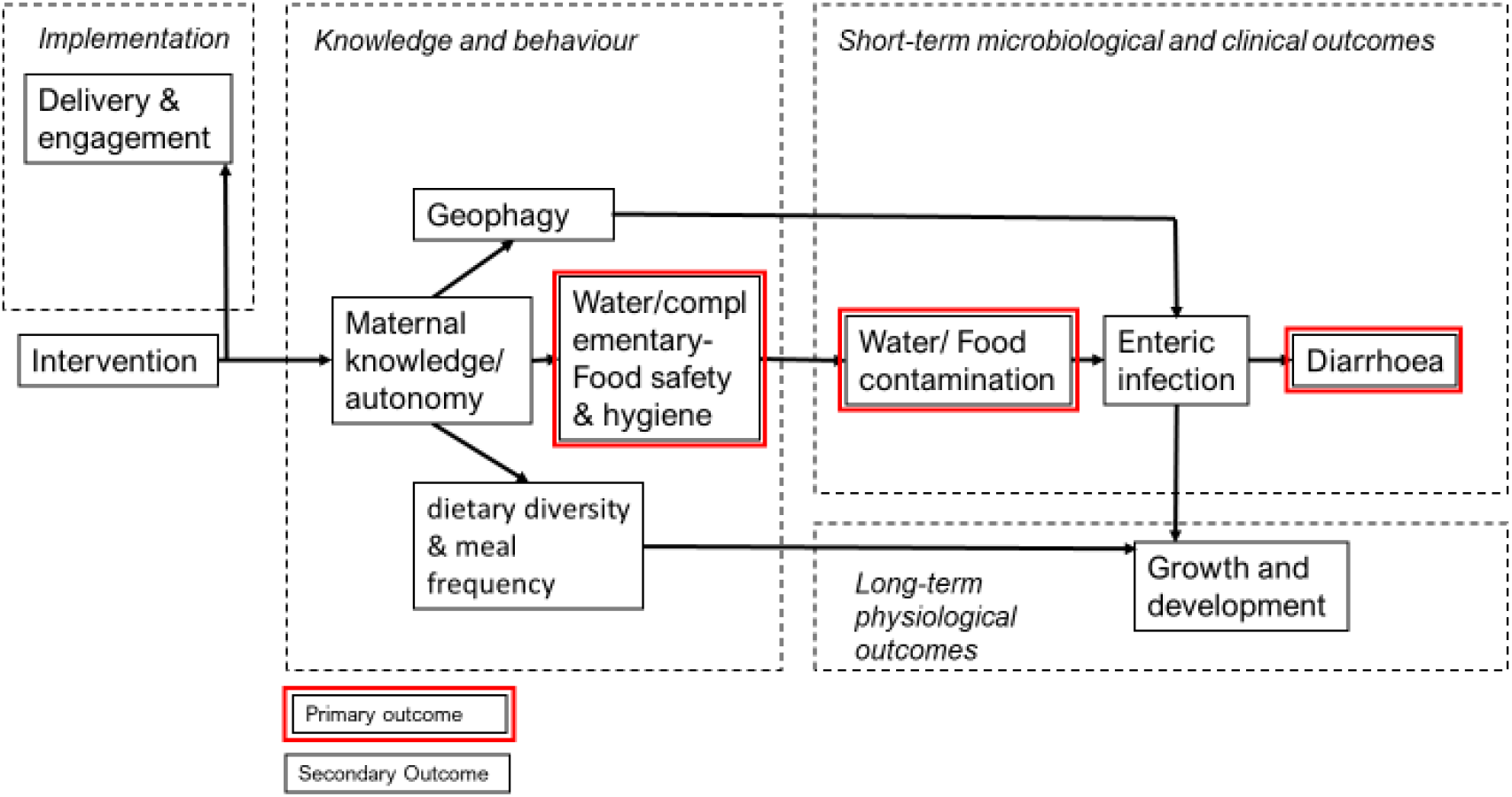
Causal diagram represented as a directed acyclic graph representing the causal assumptions of the study. The three primary outcomes in red are intended to capture the general pathway behaviour -> exposure -> incident case.

A causal effect of the intervention on parental water and food safety and hygiene behaviour is a necessary (although not sufficient) condition for the intervention to have a causal effect on food and water contamination. Our interpretation of results in this study is therefore conditional, both on the explicit causal model and assumptions, and that the interpretation of effects of the intervention on more “downstream” outcomes are conditional on effects observed on mediating outcomes. We also note that diarrhoea can be an unreliable and often very noisy outcome for trials of this nature [22], so the inclusion of outcomes on which it is causally dependent will aid to minimise the risk of faulty interpretation.

We recognise that in a larger community there would be a “virtuous circle” of lower infection rates and reduced transmission. The causal diagram therefore represents an individual-level and relatively simplified set of causal assumptions.

The remaining outcomes are considered “secondary” for the main analysis of the trial, although we will perform a full structural model analysis as a secondary analysis to compare to the main analysis to facilitate the causal interpretation of the results. Within each category of outcome we will also collect several alternative measures, although only one measure from each outcome type will be used for the main analyses described here (as listed in Table 2). A full table of all the primary and secondary outcomes is provided below (Table 2) and the alternative measures we will collect are detailed in the Supplementary Information. We also note that the knowledge and behaviour outcomes can be conceived of as “process outcomes” in alternative terminology. The trial can therefore be conceived as a hybrid implementation-effectiveness study of a complex intervention [23].

**Table 2.** List of study outcomes

| **Outcome category** | **Description** | **Method** | **Units** |
| --- | --- | --- | --- |
| **Primary outcomes** |  |  |  |
| Water and food safety and hygiene behaviour | The number of events during child's water and food preparation and handling where the mother has an opportunity to practice a pre-specified drinking-water and complementary-food safety and hygiene behaviour in which the behaviour is observed (e.g. hand washing before food preparation) of all total opportunities in the observation period | Observation | Number of opportunities met (binomial) |
| Food and water contamination | *E. Coli* count in child's food and water samples | Sample collection and field testing | Colony-forming units/gram (cfu/g) |
| Diarrhoea | Observation of stool consistency (watery diarrhoea) by field data collectors and history of at least 2 other such stools in the last 24 hours | Observation of collected stool by field worker | Dichotomous |
| **Secondary outcomes** |  |  |  |
| **Implementation** |  |  |  |
| Fidelity | Number of visits of the intervention delivered as planned | Investigator report | Number |
| Uptake | Number of the days of intervention observed by mother (Table 1) | Survey | Number |
| **Knowledge and behaviour** |  |  |  |
| Nutrition | Minimum acceptable diet based on minimum dietary diversity and minimum meal frequency they are fed during the day (DHS survey, see below) | Survey | Dichotomous |
| Geophagy | Number of behaviours observed from total opportunities in assessment of child play environment and behaviour over the observation period. | Observation | Number of opportunities met (binomial) |
| Maternal autonomy | Women's Autonomy Measure (from DHS survey). Proportion (of n=9) of household decisions the woman participates in. | Survey | Proportion |

| **Short-term microbiological and clinical outcomes** |  |  |  |
| --- | --- | --- | --- |
| Acute respiratory infection | Parental report of cough and difficulty breathing in past 7 days | Survey | Dichotomous |
| Diarrhoea hospitalisation | In-patient hospitalisation (if given a bed to stay for observation, tests or treatment for >3 hours) for diarrhoeal disease in past three months | Survey | Dichotomous |
| Enteric infection | Qualitative PCR of the following pathogens: ETEC; EAEC; EPEC; Astrovirus; Sapovirus; Rotavirus; Adenovirus; Giardia; Cryptosporidium; E. Histolytica | Sample collection and lab testing | Presence/ Absence (of each) |
| **Long-term physiological outcomes** |  |  |  |
| Physical growth | Height | On site measurement | Height for age (z-score [WHO International Growth Tables]) |
| Cognitive development | ASQ3 score for age | Survey/ observation | Number |

### Data collection procedures

#### Field data collection, food, water and stool samples

Figure 3 shows an illustration of the general data collection process for each cluster per period. Field teams will comprise 27 data collectors (one per mother-child pair), three technical assistants to do the anthropometrics and some laboratory work, and two field supervisors.

**Figure 3.**
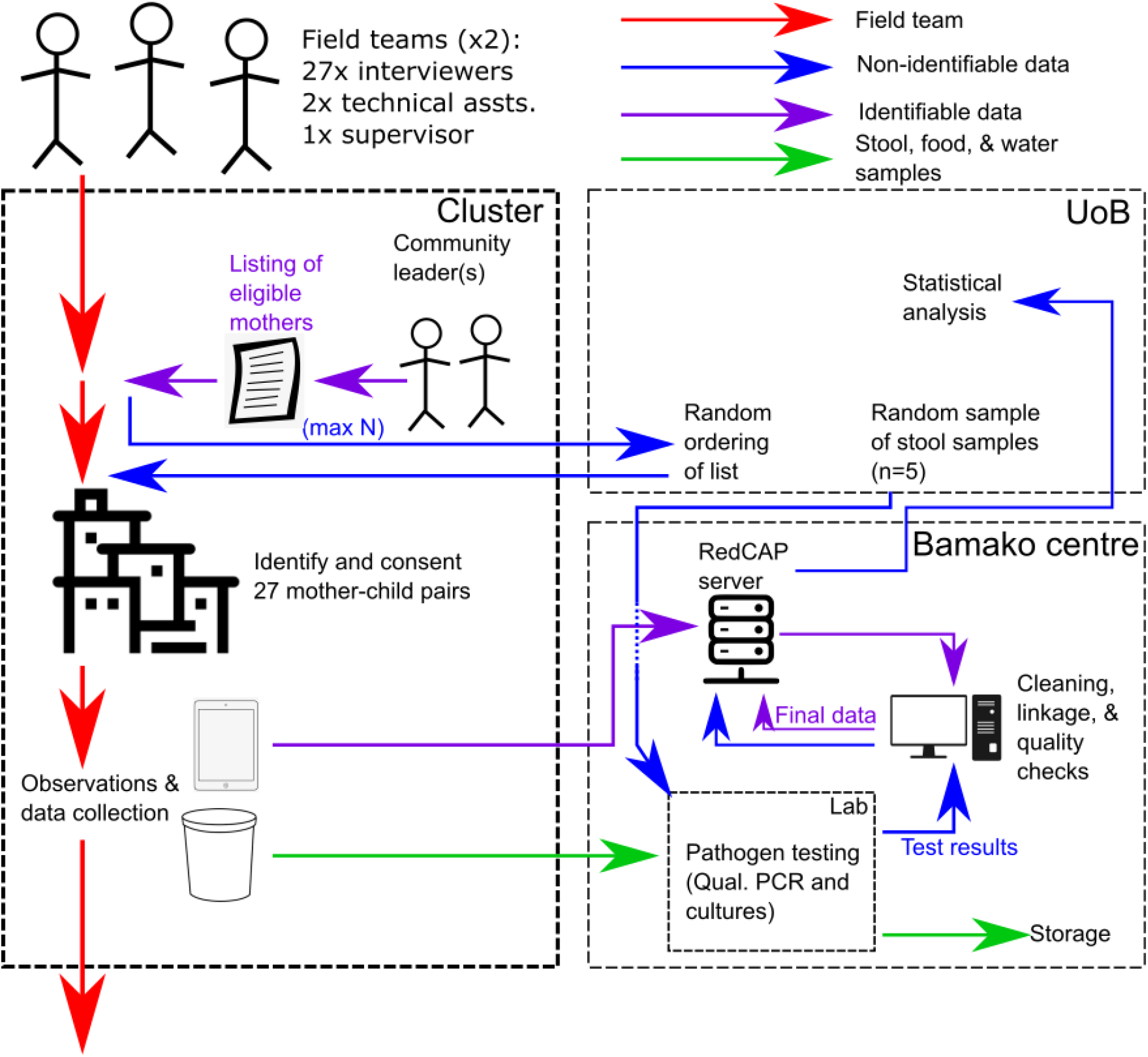
Illustrative flow diagram showing data collection procedure and data and sample flows

The day following enrolment of participants (see *Implementation*), a data collector will spend an average of 8 hours with each participating mother and child to capture the various direct observations and complete the survey questions listed in Table 2. Data capture will be on tablet device using RedCAP software; the server aggregating responses will be hosted at USTTB, Bamako. The field supervisors will conduct spot checks and sit-ins with each data collector throughout the day to check for quality and accuracy of recorded responses.

In every household, each data collector will provide the materials (cotton underwear/liner, nappies, or potty) to the household. When the child defaecates during the data collector’s 7-9 hours observation visit, a sample will be collected using aseptic techniques.

We will also collect six water and six food samples from each cluster per period. The samples will be collected from the complementary-food and water provided to the child just before consumption from the vessel or spoon or hand of the carer before it goes in the child’s mouth by the data collector using aseptic techniques. Samples can be collected from either breakfast or lunch meals. The field team will be provided in advance with a random sample of unique household identifiers for each cluster period that indicate which households and meal a sample should be taken from.

Stool samples will then be stored in liquid nitrogen capsules and water and food samples in cool boxes for transportation to the Bamako laboratory on a daily basis for analysis following completion of the data collection in the cluster. Each sampling container will have a unique predetermined anonymous ID and barcode, which will be recorded in the main data collection form.

### Lab procedures

The principal outcome at this level is the prevalence of infection of ten common pathogens in stool and E.Coli for food and water as an indicator for faecal contamination.

#### Food analysis

10gm of the sample will be weighed and suspended in a sterile vial containing 90ml of sterile Maximum Recovery Diluents (1:10 ratio). The sample will then be vortexed for 5 minutes to dislodge bacterial cells from the food particles. The homogenised sample will be allowed to stand for 10 minutes so that heavy food particles could settle before aseptically transferring 1ml of the food supernatant to the surface of the Brilliance E.coli/coliform agar. The sample collected will be spread by pressing gently with the spreader. The inoculated plates will be incubated for 24 hour +/- 1 hour at 37°C +/- 1°C. Furthermore, all the samples with over growth due to heavy coliform contaminations will be serially diluted to countable growth.

We will calculate the total number of coliforms per gram by multiplying purple and pink colonies by the dilution factor. The number of presumptive Escherichia coli will be obtained by multiplying the number of purple colonies by the dilution factor. The coliform count will be expressed in cfu/10g unit.

#### Water analysis

10ml of each water sample will be filtered using a membrane filter (0.45 μm pore size). The filtered membrane will be transferred onto an absorbent pad soaked in lauryl sulphate. The inoculated Petri dish will be labelled and incubated at 37° C for 24 hours to isolate the coliforms. We will be only counting yellow colonies using the horizontal grid lines. Coliform count will be calculated by coliform colonies multiplied by 10 (Oxfam Delagua user manual).

#### Stool Analysis

DNA will be extracted from stool samples using extraction protocols specific to the pathogens being tested for. Extracted DNA will then be mixed with diagnostic primers for the target pathogens, amplified, and run through gel electrophoresis.

The principal outcome at this level is the prevalence of infection; ten common pathogens in stool and E.Coli for food and water. Only five stools samples will be tested per cluster-period with the remainder stored for future analyses. A random sample of unique IDs will be provided to the lab in advance of the study.

#### Data processing and final dataset

Data will be downloaded and checked for consistency and errors daily when field activities are ongoing. Results from the lab procedures (see below) will be recorded on a separate form on the tablet, linked to the ID, and submitted to the server. The raw data will be identifiable as it will contain the GPS location of the household where the data collection took place alongside an identifier that is linked to the name of the participant. Only the data manager in Bamako will have access to these identifiable data. At the completion of each round of data collection, a dataset will be created combining the field and lab datasets. Individual identifiers will be anonymized and an identifiable dataset (containing GPS location) and an anonymized dataset will be generated. Access to the identifiable dataset will be limited to those requiring it for analyses approved by the MaaCiwara Management Committee. Both datasets will be made available to other researchers upon reasonable request. At the end of the trial only the final datasets will be retained.

### Analysis

#### Statistical analysis

We will conduct two types of analysis. The main analysis will follow standard recommended practice for cluster randomised trials. The main analysis will have two parts: estimation of overall treatment effects, and then a subgroup analysis. We will not use statistical significance in either reporting or for interpretation of intervention effect estimates. Our interpretations will be based on the sizes and uncertainty of treatment effect estimates conditional on the causal model we have assumed (see *Outcomes*). In particular, interpretation of an estimated treatment effect is conditional on the estimated effects of the intervention on causally antecedent outcomes. For example, if there are only small effects of the intervention on water and food safety and hygiene behaviour then any subsequent effects on food and water contamination must also be small. The conditional interpretation of our primary outcomes provides some protection against “type I errors” of interpretation and so we therefore also do not opt to adjust for multiple outcomes [24].

The secondary analysis will take a more explicit structural equation approach to account for the dependencies we assume in the data (Figure 2) and to address Objective 3, which is to estimate the effects of the intervention (or lack thereof) mediated by different potential causal pathways. However, this more complex analysis can fail (e.g. convergence failure) and so it is left as a secondary analysis. We describe each analysis in turn.

##### Primary analyses

###### Analysis 1

We will estimate overall pooled (urban and rural) intention to treat (ITT) effects for all three primary outcomes at 15 months post-intervention. We will analyse each outcome using a generalised linear mixed model. The level of observation is the individual nested within cluster-periods. Models will include an intercept, an indicator for whether the cluster had the intervention at the time, and a post-intervention time period indicator. We will also include any covariates used in the randomisation process in each model. Normally-distributed random-effects with unknown variance will be included at the cluster and cluster-period levels. Specifically, for individual *i* in cluster *j* at time *t* the outcome is *y*_*ijt*_. We specify the linear predictor as:

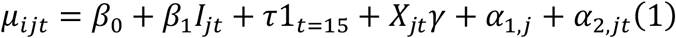

where *I*_*jt*_ is an indicator equal to one if the cluster has the treatment at time *t* and zero otherwise such that *β*_1_ is the treatment effect of interest, 1_*t*=15_ is an indicator for the post-intervention (15 month) period, *X*_*jt*_ are cluster-level covariates used in the randomisation, and 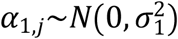 and 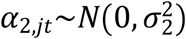 are between-cluster and within-cluster, between-period random effects, respectively. For the behavioural and diarrhoea outcomes we specify:

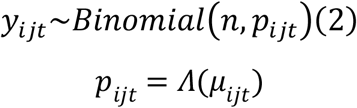

where 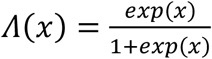 and *n* = 4 for the behaviour outcome and *n* = 1 for the diarrhoea outcome. For the contamination colony count outcome *y*_*ijt*_∼*Poisson* (*exp* (*μ*_*ijt*_)). The model assumes a cross-sectional sampling structure within cluster-periods. In the case that mothers appear in multiple rounds (with the same or different children), and we can successfully link observations, then we will treat the outcomes as repeated measures and include a mother-level random effect term.

We will report both relative effects (odds ratios for binomial models and rate ratios for Poisson models) and absolute differences for all outcomes (primary outcomes, secondary outcomes in Table 2 and alternative outcomes in Table S1, Supplementary File 3) along with 95% confidence intervals and the p-value associated with a two-sided test of no difference. For non-linear models the absolute differences will be averaged over the study population (the average marginal effect) using a marginal standardisation approach. We will also report estimated within- and between-cluster variance.

###### Analysis 2 (Subgroup analyses)

We will conduct two subgroup analyses. For the first we aim to compare the effect of the intervention in urban and rural settings. We modify the models described above by adding an indicator for urban clusters and its interaction with the treatment indicator:

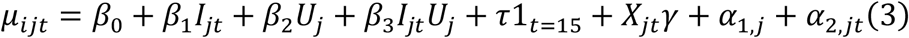

We will report the parameter *β*_3_, which can be interpreted as the ratio of odds ratios or ratio of rate ratios between urban and rural settings, along with 95% confidence intervals. We will also report the estimated effect sizes in both relative and absolute terms for urban and rural clusters. The second subgroup analysis will compare 4- and 15-month post-intervention periods to evaluate potential decay or strengthening of effects over time. For this model we will use all three periods of data. The mean function will be as specified in Equation (3) but with an indicator for the 4-month period (i.e. 1_*t*=4_) instead of *U*_*j*_. We will report the same subgroup effects and comparisons as for the urban and rural comparison.

##### Secondary analyses

###### Structural model

Our secondary analysis will take an exploratory and explicitly structural approach, based on the assumed causal relations specified in Figure 2 to facilitate interpretation of the causal effects of the intervention. For this analysis we will use Bayesian methods, given the complexity of the model structure, which is more amenable to Markov Chain Monte Carlo (MCMC) methods. We will use the primary and secondary outcomes, each of which will form a sub-model of the overall model. An individual level random effect will be included, in addition to a cluster-level effect, which will appear in each sub-model weighted by a factor loading. These random effects will allow for correlation between the outcomes at an individual and cluster level. We will use weakly informative priors for the model parameters and hyperparameters. The structural model will enable us to extract several effects of interest. In particular, the effects of the intervention on enteric infection and growth as mediated by the different pathways in the diagram [25]. One comparison of particular interest is whether the effects of the intervention differ in urban and rural areas, thus this analysis will take place separately for urban and rural clusters.

### Sample size and power

We plan to use stratified randomisation to allocate 120 communities to two arms and recruit 3,240 mother-child pairs in total at baseline (27 per cluster-period), midline (4-months post-intervention), and endline (15 months post-intervention) as three cross-sectional data collection rounds. For the three outcomes we will collect:

1. Water and Food safety and hygiene behaviour: a binomial outcome with four questions per pair for all 27 mother-child pairs.
2. Food and water contamination: 10 samples randomly selected from the 27 total samples (1 per pair) will be analysed.
3. Diarrhoea: A single dichotomous observation for each of the 27 mother-child pairs.

We report the power for Analyses 1 and 2 for the three primary outcomes listed in Table 3 for a range of effect sizes assuming an intraclass correlation coefficient of 0.02, a cluster autocorrelation coefficient of 0.8, and a type I error rate of 5%. Generation of ICC and CAC estimates follow recommended guidance [26], and we have used values informed by similar outcomes in similar settings for the ICC (noting that ICCs for process outcomes tend to be higher for those for clinical outcomes [27]. For the CAC where we have limited information on values we have used the values of 0.8 and 0.9 as recommended in the literature [28]. We consider sensitivity to a range of plausible values: power calculations with alternative values of the ICC and CAC are provided in the Supplementary File 3. For Analysis 1 we used the method described by Hemming *et al* [29] and for the sub-group analyses we adapted this method to that reported by Demidenko [30]. We did not include covariate effects nor the randomisation procedure in the power calculations, thus these results can be considered conservative.

**Table 3.**
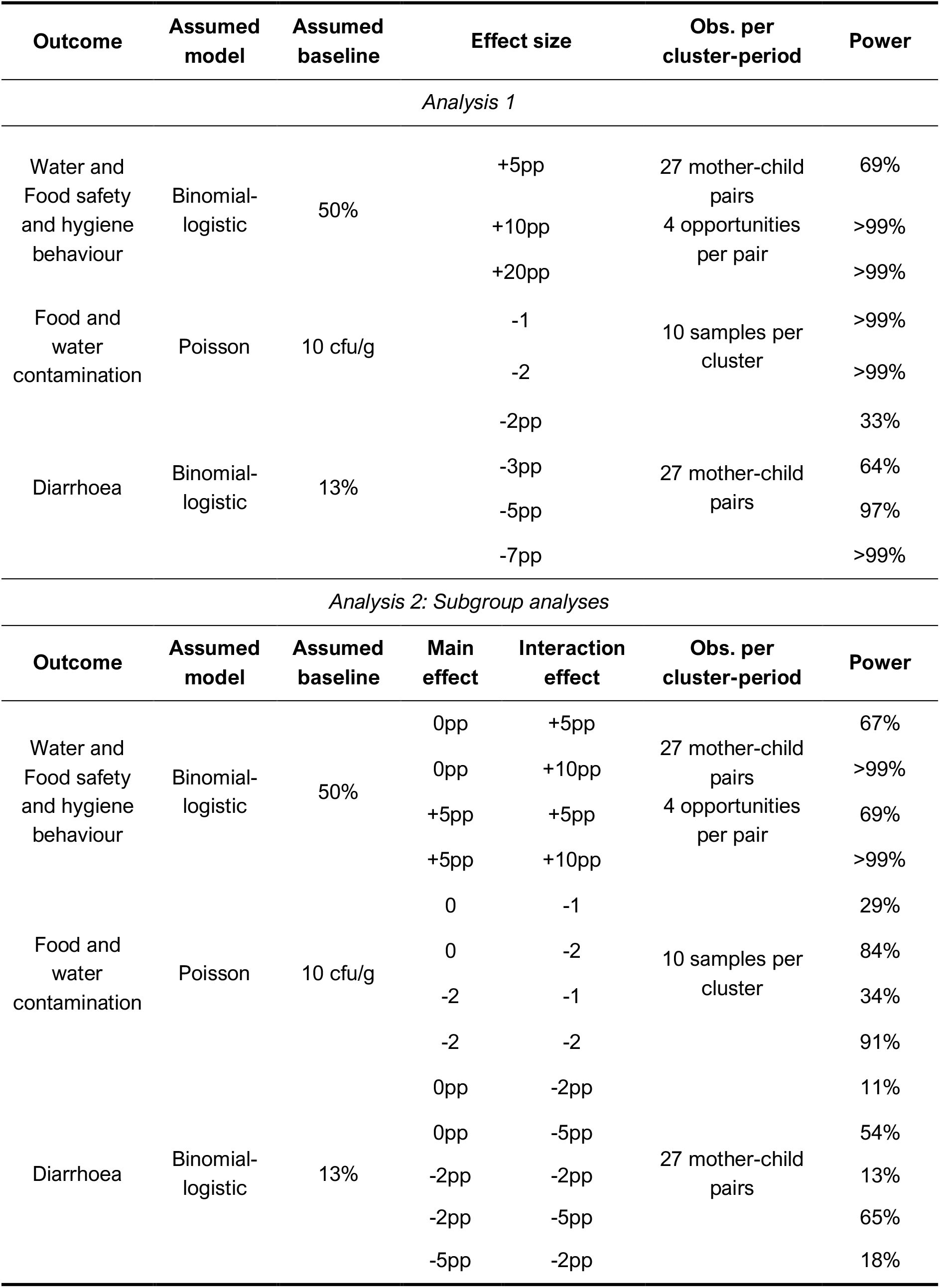
Power for the three main outcomes for different combinations of effect sizes and assuming a ICC of 0.02, a CAC of 0.8, and a type I error rate of 0.05. Power for non-linear models calculated using a normal approximation. pp = percentage point

#### Qualitative analysis

In parallel to the trial, comparative focussed ethnographic research will be carried out in a random sub-sample of the control (2x rural, 2x urban) and intervention (3x rural and 3x urban) clusters, with a focus on understanding the social, political and cultural context in which the intervention is being implemented and the acceptability of the intervention in this context, particularly the local arts-based campaign approach. The qualitative analysis will enable a more nuanced interpretation of issues of potential scalability and transferability to different cultural contexts, through refining the programme’s theory of change.

The primary method will be participant-observation, via key informants (local artists and families in the selected clusters). The ethnographic fieldworkers will be independent of the intervention delivery and trial outcomes measurement teams and will be clear with participants that they have separate objectives. Fieldnotes will be taken using proformas: handwritten notes will be taken in the field, then typed up to create a full set of fieldnotes within 24 hours of leaving the field. Fieldnotes will be cross-checked during debrief with other team members and clarifications and reflections added. Supplementary visual methods (photographs, sociograms) and ethnographic (unstructured) interviews will be used when needed to offer further insights or explanations of observed events. Data from these methods will be incorporated into fieldnotes.

In addition, after completion of the trial, up to 12 focus groups will be conducted with mothers, fathers, grandmothers and village elders in a sub-sample of intervention clusters (different to the ones selected for the ethnography sub-analysis) to draw out their reflections on the intervention process and impact.

Data analysis will take place alongside data collection guided by a realist theoretical approach, with a focus on understanding the influence of context on the mechanisms of action in the intervention. This may include themes such as (i) the influence of urban and rural context upon intervention adaptation and implementation, (ii) of the role of cultural (African) dramatic arts beyond information transfer to a mediator for behaviour change, (iii) of mother’s empowerment, (iv) implementation processes and (v) social network/support mapping.

We will be working across numerous languages (local dialects, French, English) and, therefore, we will use the Framework method for the management and analysis of qualitative data. This method is appropriate in this context because it requires the production of summarized data (field-notes and/or transcriptions), which can then be translated into French and English and then compared and contrasted more easily between themes and sites.

#### Health economic analysis

There is very little economic evidence about sanitation and hygiene programmes in general [31] or on the cost-effectiveness of particular types of intervention [32]. The primary base case analysis will adopt a societal perspective as far as possible. Resource use data will be collected prospectively to estimate the costs in each of the trial arms [33]. This will include costs incurred by the agencies responsible for setting up and delivering the intervention and for the households in the intervention and control villages. Costs associated with developing and delivering the intervention will be collected via trial reporting mechanisms. Costs to the household associated with episodes of childhood diarrhoea and other illness will be captured via a questionnaire which will include healthcare resource use, impacts on household productivity and other costs. The questionnaire will be delivered to participants in the trial during two follow up visits. We will use established methods to estimate the costs associated with caring for an ill child [34]. The potential costs for the participants associated with the behaviour change promoted will also be examined.

We will evaluate whether the intervention is cost-effective by extending from the framework proposed by Borghi et al [35]. For the base case we will assess cost-effectiveness based on the primary trial outcome and assess the incremental cost per case of diarrhoea avoided at 15-months post-intervention. Where possible, we will aim to collect data to enable a broader analysis of costs and benefits (for other health outcomes and beyond health). A range of sensitivity analyses will be undertaken to explore the impact of uncertainty on the study results [36].

### Ethics

#### Research Ethics Approval

The study received full ethical approval from the University of Birmingham Science, Technology, Engineering, and Mathematics (STEM) Research Ethics Committee (ERN_20-0625). Also, in Mali, full ethical approval was granted by the Faculty of Medicine, and stomatology, and Pharmacology, University of Science, Techniques, and Technologies of Bamako (Letter Number 2020/253/CE/FMOS/FAPH).

#### Protocol Amendments

Amendments to the protocols can only be made by the chief and/or principal investigators or their designees, with agreement from the sponsors. Substantial amendments will also be reviewed by the project’s oversight committees, stakeholders, and must be approved by the Malian ethics committee. Non-substantial or administrative amendments will be documented but do not require formal approval.

#### Data Monitoring Committee

We will not specify any “stopping rules” for the trial as the intervention is community-based, non-invasive, and non-clinical and presents no risk of harm to the study participants. The role of the Data Monitoring Committee is therefore to monitor trial conduct and data quality rather than identify safety events. The Trial Steering Committee will also constitute the Data Monitoring Committee for the trial and so will adopt its responsibilities, and is comprised of independent experts in the area of WASH interventions and their evaluation. The Steering Committee will meet six times over the three years the trial is taking place. Prior to trial commencement it will advise on design of the study and analysis. Following completion of baseline data collection the committee will review a summary of data collected and data quality indicators to be specified. The trial statistician in Bamako will prepare this report and analysis. The Committee will also meet to repeat the review following completion of each subsequent wave of the study.

#### Community Engagement and Involvement

To improve safety and cultural appropriateness of this trial, lay individuals from target populations will be substantially involved in the research, through formative research and throughout the study in the following ways:

1. Community leaders and mothers in communities were and will continue to be consulted during the development of the study, in both rural and urban Mali.
2. Formative research took place through running focus groups and home visits with community members, such as mothers, fathers, grandmothers and community leaders. The working group that will adapt the intervention will include non-researchers who are themselves from communities: a creative team with local traditional communicators and dramatic artists and public health officers working in the existing Mali community health system. This allows for adaptation of the intervention and the survey instruments to the community, allowing for the study to be as acceptable and relevant as possible.
3. The trial’s Mali Expert Advisory Group will include two community members to provide ongoing support and advise on the management of research, undertaking of research, and analysis and interpretation of results.4. An extensive list of stakeholders and policy makers (and community members at the end of the study) will also receive update newsletters and the results of the study through the Communication and Dissemination Plan, encouraging them to feedback and send views and questions. The trial website will also allow for such interactions through different contact routes in Mali and UK.

## Data Availability

Both datasets will be made available to other researchers upon reasonable request. At the end of the trial only the final datasets will be retained.

## Funding

Medical Research Council (MRC), UK Research and Innovation (UKRI) Global Challenges Research Fund (GCRF) MR/T030011/1. RJL and SMH are also supported by the National Institute for Health Research (NIHR) Applied Research Collaboration (ARC) West Midlands. The views expressed are those of the author and not necessarily those of the NIHR or the Department of Health and Social Care

## Conflicts of interest

None declared

## Author contributions

SM-H, RJL, MSI, SC, CS, YD, KJ, KK, OK, and OT conceived the project and developed the original project plan. SW drafted the first version of this protocol and reviewed and revised the methodology. SIW, KH, and RJL developed the statistical methods. SIW and EA revised subsequent versions of the protocol. All authors reviewed and approved the final version of the protocol.

## Trial sponsor

University of Sciences, Techniques and Technologies of Bamako (USTTB) at the University of Bamako, Bamako, Mali.

## Version History

1.1 07 Dec 2021
1.2 31 Mar 2022 – *Amended funding statement to include support from the National Institute for Health Research (NIHR) Applied Research Collaboration (ARC) West Midlands.*

## Notes

### Competing Interest Statement

The authors have declared no competing interest.

### Clinical Trial

ISRCTN14390796

### Funding Statement

This study was funded by Medical Research Council (MRC), UK Research and Innovation (UKRI) Global Challenges Research Fund (GCRF) MR/T030011/1. RJL and SMH are also supported by the National Institute for Health Research (NIHR) Applied Research Collaboration (ARC) West Midlands. The views expressed are those of the author and not necessarily those of the NIHR or the Department of Health and Social Care

### Summary of Updates

Amended funding statement to include support from the National Institute for Health Research (NIHR) Applied Research Collaboration (ARC) West Midlands.

## REFERENCES

1. (WHO), W.H.O. Diarrhoea Disease. 2017; Available from: https://www.who.int/news-room/fact-sheets/detail/diarrhoeal-disease.

2. Troeger, C., et al., Estimates of global, regional, and national morbidity, mortality, and aetiologies of diarrhoeal diseases: a systematic analysis for the Global Burden of Disease Study 2015. The Lancet Infectious Diseases, 2017. 17(9): p. 909–948.

3. Checkley, W., et al., Multi-country analysis of the effects of diarrhoea on childhood stunting. Int J Epidemiol, 2008. 37(4): p. 816–30.

4. Walker, C.L.F., et al., Does childhood diarrhea influence cognition beyond the diarrhea-stunting pathway? PloS one, 2012. 7(10): p. e47908.

5. Walker, C.L., et al., Global burden of childhood pneumonia and diarrhoea. Lancet, 2013. 381(9875): p. 1405–16.

6. Grace, D., Food Safety in Low and Middle Income Countries. International Journal of Environmental Research and Public Health, 2015. 12(9): p. 10490.

7. Hulebak, K.L., Hazard Analysis and Critical Control Point (HACCP) History and Conceptual Overview. Risk Analysis: An International Journal, 2002. 22(3): p. 547–553.

8. World Health Organization (WHO), Application of the hazard analysis critical control point (HACCP) system for the improvement of food safety., in WHO supported case studies on food prepared in homes, at street vending operations and in cottage industries. 1993, WHO.

9. George, C.M., et al., Geophagy is associated with environmental enteropathy and stunting in children in rural Bangladesh. The American journal of tropical medicine and hygiene, 2015. 92(6): p. 1117–1124.

10. Islam, M.S., et al., Hygiene intervention reduces contamination of weaning food in Bangladesh. Trop Med Int Health, 2013. 18(3): p. 250–8.

11. Toure, O., et al., Piloting an intervention to improve microbiological food safety in Peri-Urban Mali. Int J Hyg Environ Health, 2013. 216(2): p. 138–45.

12. Gautam, O.P., et al., Trial of a novel intervention to improve multiple food hygiene behaviours in Nepal. American Journal of Tropical Medicine & Hygiene, 2017. Volume 96(Issue 6): p. 1415–1426

13. Tylleskär, T., et al., Exclusive breastfeeding promotion by peer counsellors in sub-Saharan Africa (PROMISE-EBF): a cluster-randomised trial. The Lancet, 2011. 378(9789): p. 420–427.

14. Perkins, J.M., S.V. Subramanian, and N.A. Christakis, Social networks and health: a systematic review of sociocentric network studies in low-and middle-income countries. Soc Sci Med, 2015. 125: p. 60–78.

15. Pleasant, A., et al., A qualitative first look at the Arts for Behavior Change Program: Theater for Health. Arts & Health, 2015. 7(1): p. 54–64.

16. Manaseki-Holland, S., et al., Effects on childhood infections of promoting safe and hygienic complementary-food handling practices through a community-based programme: A cluster randomised controlled trial in a rural area of The Gambia. PLoS medicine, 2021. 18(1): p. e1003260.

17. Manjang B, Investigating effectiveness of behaviour change intervention in improving mothers weaning food handling practices: A cluster randomised controlled trial in rural Gambia, in Institute of Applied Health Research. 2017, University of Birmingham: Birmingham.

18. Ezeh, A., et al., The history, geography, and sociology of slums and the health problems of people who live in slums. The lancet, 2017. 389(10068): p. 547–558.

19. Prost, A., et al., Women’s groups practising participatory learning and action to improve maternal and newborn health in low-resource settings: a systematic review and meta-analysis. The Lancet, 2013. 381(9879): p. 1736–1746.

20. Nakahara, S., et al., Differential effects of out-of-home day care in improving child nutrition and augmenting maternal income among those with and without childcare support: a prospective before-after comparison study in Pokhara, Nepal. 2010. 97(1): p. 16–25.

21. Hoffmann, T.C., et al., Better reporting of interventions: template for intervention description and replication (TIDieR) checklist and guide. Bmj, 2014. 348.

22. Rego, R., Watson, SI, Ul-Alam, A, Abdullah, SA, Yunus, Alam, IT, Chowdhury, ASMHK, Haider, SMA, Faruque, ASG, Khan, AI Hofer, T, Gill, P, Islam, MS, Lilford, RJ., A Comparison of Traditional Diarrhoea Measurement Methods with Microbiological and Biochemical Indicators in the Cox’s Bazar Displaced Persons Camp: A Cross-Sectional Observational Study.. EClinicalMedicine 2021.

23. Wolfenden, L., et al., Designing and undertaking randomised implementation trials: guide for researchers. bmj, 2021. 372.

24. US Food and Drug Administration, Multiple Endpoints in Clinical Trials. Guidance for Industry. tech. rep.,, US Food and Drug Administration, Editor. 2017: Silver Spring, MD, USA.

25. Watson, S.I., et al., Estimating the effect of health service delivery interventions on patient length of stay: A Bayesian survival analysis approach. Journal of the Royal Statistical Society: Series C (Applied Statistics), 2021.

26. Eldridge, S.M., et al., How big should the pilot study for my cluster randomised trial be? Statistical methods in medical research, 2016. 25(3): p. 1039–1056.

27. Campbell, M.K., P.M. Fayers, and J.M. Grimshaw, Determinants of the intracluster correlation coefficient in cluster randomized trials: the case of implementation research. Clinical Trials, 2005. 2(2): p. 99–107.

28. Hooper, R. and L. Bourke, Cluster randomised trials with repeated cross sections: alternatives to parallel group designs. bmj, 2015. 350.

29. Hemming, K., et al., A tutorial on sample size calculation for multiple-period cluster randomized parallel, cross-over and stepped-wedge trials using the Shiny CRT Calculator. International journal of epidemiology, 2020. 49(3): p. 979–995.

30. Demidenko, E., Sample size and optimal design for logistic regression with binary interaction. Statistics in medicine, 2008. 27(1): p. 36–46.

31. Hutton, G. and M. Varughese, The costs of meeting the 2030 sustainable development goal targets on drinking water, sanitation, and hygiene. 2016.

32. Hutton, G. and C. Chase, The knowledge base for achieving the sustainable development goal targets on water supply, sanitation and hygiene. International journal of environmental research and public health, 2016. 13(6): p. 536.

33. Drummond, M.F., et al., Methods for the economic evaluation of health care programmes. 2015: Oxford university press.

34. Whittington, D. and J. Cook, Valuing changes in time use in low-and middle-income countries. Journal of Benefit-Cost Analysis, 2019. 10(S1): p. 51–72.

35. Borghi, J., et al., Is hygiene promotion cost-effective? A case study in Burkina Faso. Tropical Medicine & International Health, 2002. 7(11): p. 960–969.

36. Glick, H.A., et al., Economic evaluation in clinical trials. 2014: OUP Oxford.

