## Supplementary File 2 for "Protocol for a cluster randomised trial to evaluate a community-level complementary-food safety and hygiene and nutrition intervention in Mali: The MaaCiwara study"

### Community Leader Information and Consent Form

Date ....../....... /.......

**Ethics Committee approval number**: **2020/253/CE/FMOS/FAPH**

**Title of Study: Formative Research**

**Sponsors**: University of Science, Technology and Engineering of Bamako, University of Birmingham

**What is informed consent?**

As the leader of your community, we have come to visit you and get your permission because your community is being invited to participate in a research study that aims to improve the health and lives of young children and their families. Participating in a research study is not the same as receiving regular medical care. The goal of regular medical care is to improve a person's health. The purpose of a research study is to gather information to help improve the health of others, and you or your family may benefit from it as well. But it is your choice whether or not to participate.

Before you decide, you need to understand all the information about this study and what it will mean for your community. Please take the time to read the following information or have the information explained to you in your own language. Listen carefully and don't be afraid to ask if there is anything you don't understand. Ask to have it explained to you until you are satisfied. You can also consult with your family, family members or others before deciding to participate in the study.

If you decide that your community will participate in the study, you will be asked to sign or thumbprint a consent form indicating that you agree to participate in the study.

**Why is this study being conducted?**

This study is intended to help us understand how we can improve the health of children aged 6 to 24 months in rural and urban Mali and to test a new program that we believe will achieve this goal. We are particularly interested in improving family practices around food hygiene, nutrition and safe play for young children. We involve children aged 6 to 36 months because children's health at this stage can have an impact on health for the rest of their lives. We've done this research in The Gambia in a small number of villages and found that it helps children to be healthier, families happier and communities more united. We want to know if it will have the same benefits in Mali and if we should offer it to more communities.

**If you participate, what will happen? (What does this study mean to you?)**

We chose 120 communities in Mali in Bamako, Koulikoro, Sikasso and Segou and the villages in the rural areas around these cities. We chose the communities using a random method and we are delighted that by chance your village/community is one of them.

We want to see how we can improve the food content and food safety/hygiene for baby food in families with children aged 6-24 months, and how we can improve the way these children play so that they don't have unhealthy things when they play. This will be done through a community celebration and campaign program with community events led by traditional singers and drummers. To test whether the program works or not, half of the villages/communities will receive the program, and the other half will receive a different, simpler program. We will conduct surveys and compare the two different programs and study their benefits.

The main program will be a team of 3-4 public health workers and 2 singers/drummers who will visit your village/community on 5 different days for 6 weeks (days 1, 2, 17, 25, 40). They will make street announcements, put up posters to educate mothers, visit mothers in their homes and invite mothers and other community members to community events. Community events will include plays, songs, music, contests, videos and support from older mothers. We need you and your wife's support during these events to encourage community participation.

We will also ask your community to nominate one or two older mother volunteers to support and encourage the young mothers in our program and we will train them for this work with a small incentive/donation to encourage them.

The other program includes a one-day visit to your village/community to invite community members and mothers for a day of teaching about water use in gardens and home yards.

At this time, we do not yet know what program will be offered to your village. Once you agree to participate, we will be able to inform you about the program that will be offered to your community/village. However, if you do not receive the 5-day visit program, we will make sure that your community is informed at the end of our work in 18 months.

In addition to these health information and campaign programs to measure whether our work is beneficial or not and whether we should do these programs in other communities, we will also come for 3 separate days to your village to visit the homes of 30 mothers and collect survey data on their daily work and childcare practices to see if our work has been helpful or not. The 3 days will take place before the program, after 4 months and after 18 months. During the 18 months, three researchers may visit your village or community for a few days to ask mothers and other community members what they think of the program, informally or otherwise. For all these surveys or discussions, we will get permission/consent from the people we talk to (participants), such as mothers and their families, and we will only do so if they agree.

What happens next: If you agree to participate, we will come to your community in about 1 month to collect data from the first home visiting survey of 30 mothers of young children. Then we will randomly select the program your village will receive and tell you when we can visit your community for the program.

**What will we do with the information we collect?**

All information collected will be treated as confidential and will not be used for any purpose other than research. We will create codes for each of the communities/villages and families so that the names of the individuals and the name of the village/community will not appear in our files or in any documents. This information will be kept secure so that only research staff can see it and they will keep all information very confidential. We may need this data in the future to look at other research questions, but if we do, we will remove all names of people and all village/community information.

**What harm or discomfort can you expect from the study?**

There is no harm or discomfort associated with this study.

**What benefits can you expect from this study? Will you be compensated for your participation in the study?**

This study means that you and your village/community will benefit from healthier and better nourished children and families. You will also help other communities in Mali because the results of the study could help the government to improve health and social services in Mali and improve the lives and health of children. Other African countries will also learn and benefit from what we learn from this research, as many countries in West Africa have the same health problems with their children.

Also, to thank the mothers for their time, we will give them a small gift.

**What happens if you refuse to participate in the study or change your mind later?**

You are free to participate or not in the study and nothing else will happen. You can also withdraw your consent to participate from your community at any time. In addition, mothers have the right to withdraw their information up to one week after our study and home visit without giving a reason, while we can still remove your information from our records.

**Who should you contact if you have any questions?**

If you have any questions or concerns, you can contact 66 72 90 13 or 76 32 96 04 and you can always call the personal numbers of the study staff that have been given to you. Please feel free to ask any questions you may have about the study.

**Who reviewed this study?**

This study has been reviewed and approved by the Ethics Committee of the Faculty of Medicine and Odonto-stomatology of the USTTB of Mali and the University of Birmingham's Ethical Review Committee, which is made up of scientists and ordinary people to ensure that the study protects your rights and welfare.

**Community Leader Consent Form**

**Declaration of fair treatment**

This information is being collected as part of a research project to gather information on child health in Mali by the Institute of Applied Health Research, University of Birmingham, in collaboration with the University of Science, Technology and Engineering, Bamako. The information you provide and that which may be collected as part of the research project will be entered into a filing system or database and will only be accessible to authorised personnel involved in the project. The information will be held by the University of Birmingham and will only be used for research, statistical and audit purposes. By providing this information, you consent to the University retaining it for the purposes stated above. The information will be processed by the University of Birmingham in accordance with the provisions of the Data Protection Act 1998. No personally identifiable data will be published.

**Statements of understanding/consent**

- I confirm that I have read and understood the information sheet for participants in this study. I had the opportunity to ask questions when necessary and received satisfactory answers.
- I understand that participation in my community is voluntary and that I am free to withdraw at any time without giving a reason. If I withdraw, my data will be removed from the study and destroyed.
- I understand that my personal data will be processed for the purposes detailed above in accordance with the Data Protection Act 1998.
- In view of the above, I agree to have my community participate in this study.

**Name, signature and date**

___________________________________________________________________________

Name of participant :

_______________________________________________	______________

Signature or left thumbprint of participant	Date

_______________________________________________	______________

Name of researcher / person obtaining consentDate

### Information and Consent Form for Mothers

Date ....../....... /.......

**Ethics Committee approval number**: **2020/253/EC/WOSF/APHF**

**Title of Study: Formative Research**

**Sponsors**: University of Science, Technology and Engineering of Bamako, University of Birmingham

**What is informed consent?**

You are being asked to participate in a research study. Participating in a research study is not the same as receiving medical care. The purpose of medical care is to improve health. The purpose of a research study is to gather information to help improve the health of others, and you or your family can benefit from it too. But it is your choice whether or not to participate.

Before you decide, you need to understand all the information about this study and what it will involve. Please take the time to read the following information or have it explained to you in your language. Listen carefully and do not hesitate to ask if there is anything you do not understand. Ask for it to be explained to you until you are satisfied. You may also want to consult with family members or others before deciding to participate in the study.

If you decide to participate in the study, you will be asked to sign or fingerprint a consent form indicating that you agree to participate in the study.

**Why is this study being conducted?**

The purpose of this study is to help us understand how children aged 6 to 36 months and their mothers spend their days in rural and urban Mali. This information will help the government and service providers deliver better health and social services to communities like yours. We are interested in mothers and children aged 6 to 36 months because the activities of the mother are important to the whole family and the health of children at this stage can influence the rest of their lives.

**What would happen if you agreed to participate in this study? (What does this study mean to you?)**

We are interested in your activities during the day, the health and history of the baby, mother, and family. For this study, I (investigator) will stay with you (mother/other caregiver) from the time you wake up in the morning until after lunch (investigator asks for wake-up time and writes here ---- am). During this time, I will do the following and take some notes on my device/tablet or on paper:

- watch you do your daily work,
- watch the baby's activities,
- ask questions about your daily work at home, your baby's health, other general information about the baby, you (mother) and your family,
- measure your child's height, weight and arm circumference,
- Take a small sample of food (1 tablespoon) and water for testing,
- Take a sample of your baby's stool for analysis,

So after your consent this evening, I will have, if I may, some general questions about your family and your life in general.

**What will be done with the information we collect?**

All information collected will be treated as confidential and will not be used for any purpose other than research. We will create codes for each of the communities/villages and families so that your name and the name of your village will not appear in our records. This information will be kept in a secure place so that only research staff can see it. We may need to share the data later so that we can look at other things, but if we do, we will remove all identifying information.

**What are the disadvantages of participating in this study? (What harm or discomfort can you expect in the study?)**

There is no harm or discomfort associated with this study. I will only stay with you all day. I (investigator) will bring whatever I need (my own lunch/food) so as not to inconvenience you. We would not like you to treat me as a guest but to ignore my presence in your home as much as possible. We would like you to carry out your daily activities in the usual way.

**What benefits can you expect from the study? Will you be compensated for your participation in the study?**

The results of the study could help the government improve health and social services in Mali. This means that you and your family will also benefit in the long run.

To thank you for your time and to compensate for the small amount of food we take, you will receive a small jar as a gift.

**What happens if you refuse to participate in the study or change your mind later?**

You are free to participate or not in the study and nothing else will happen. You also have the right to request the deletion of your data up to one week after my visit to your home, during which time we may still delete your data from our records, without giving any reason for doing so. If you decide to withdraw your data during the course of the study, we will not process the information collected from you and this information will be deleted from our records.

**Who should you contact if you have any questions?**

If you have any questions or concerns, you can contact 66 72 90 13 or 76 32 96 04 and you can always call the personal numbers of the research team members given to you. Please feel free to ask any questions you may have about the study.

**Who reviewed this study?**

This study has been reviewed and approved by the Ethics Committee of the Faculty of Medicine and Odontology of the USTTB of Mali and the Ethics Committee of the University of Birmingham, which is made up of scientists and ordinary people to ensure that the study protects your rights and welfare.

**Consent form for participating mothers**

**Declaration of fair treatment**

This information is being collected as part of a research project on children's health in Mali by the Institute of Applied Health Research at the University of Birmingham, in collaboration with the University of Science, Technology and Engineering in Bamako. The information you provide and that which may be collected as part of the research project will be entered into a file or database and will only be accessible to authorised personnel involved in the project. The information will be held by the University of Birmingham and will only be used for research, statistical and audit purposes. By providing this information, you consent to the University storing your information for the purposes stated above. The information will be processed by the University of Birmingham in accordance with the provisions of the Data Protection Act 1998. No personally identifiable data will be published.

**Statements of agreement / consent**

- I confirm that I have read and understood the participant information leaflet for this study. I have had the opportunity to ask questions if necessary and have received satisfactory answers.

- I understand that my participation is voluntary and that I am free to withdraw at any time without giving a reason. If I withdraw my data will be deleted from the study and destroyed.

- I understand that my personal data will be processed for the purposes detailed above in accordance with the Data Protection Act 1998.

- On the basis of the above, I agree to participate in this study.

**Name, signature and date**

___________________________________________________________________________

Name of participant :

_______________________________________________	______________

Signature or left thumbprint of participant	Date

_______________________________________________	______________

Name of researcher / person obtaining consentDate
