## Supplementary File 3 for "Protocol for a cluster randomised trial to evaluate a community-level complementary-food safety and hygiene and nutrition intervention in Mali: The MaaCiwara study"

**SUPPLEMENTARY TABLES**

**Table S1** Alternative measures of outcomes collected in the study

| **Outcome category** | **Description** | **Method** | **Units** |
| --- | --- | --- | --- |
| **Alternative primary outcomes** | | | |
| Water and Food safety and hygiene behaviour | Availability and use of soap in key areas for handwashing | Observation | Dichotomous |
| Food and water contamination | *E. Coli* count in family/adult food | Sample collection and field testing | cfu/g |
| Diarrhoea | 7-day parental report of child diarrhoea episodes (3 or more watery stools in 24hrs) | Survey | Dichotomous |
| **Alternative secondary outcomes** | | | |
| ***Implementation*** | | | |
| Fidelity | N/A |  |  |
| Uptake | N/A |  |  |
| ***Knowledge and behaviour*** | | | |
| Nutrition | Minimum dietary diversity of 5 out of 8 food groups: Breastmilk; Grains, roots and tubers; Legumes and nuts; Dairy products (infant formula, milk, yogurt, cheese); Flesh foods (meat, fish, poultry and liver/organ meats); Eggs; Vitamin A rich fruits and vegetables; Other fruits and vegetables | Survey | Dichotomous |
|  | A minimum meal frequency of: 2 or more solid or semi-solid or soft feeds for breastfeeding children age 6-8 months, or 3 or more solid or semi-solid or soft feeds for breastfeeding children age 9-23 months; or 4 or more solid or semi-solid or soft or milk feeds for non-breastfeeding children age 6-23 months where at least one of the feeds must be a solid, semi-solid, or soft feed. | Survey | Dichotomous |
| Geophagy | Accessibility of play area to animals. | Survey | Dichotomous |
| Maternal knowledge and autonomy | Proportion of women who have achieved each stage of the mothers’ MaaCiwara intervention competition after pledging on the 5^th^ visit. | Survey | Proportion |
| ***Short-term microbiological and clinical outcomes*** | | | |
| Acute respiratory infection | In-patient hospitalisation (if given a bed to stay for observation, tests or treatment for >3 hours) for respiratory illness in past three months | Survey | Dichotomous |
| Enteric infection | N/A |  |  |
| **Long-term physiological outcomes** | | | |
| Physical growth | Mid-upper arm circumference (MUAC) | On site measurement | Number |
|  | Weight | On site measurement | Weight for age (z-score [WHO International Growth Tables]) |
| Cognitive development | N/A |  |  |

**Table S2** Power for the three main outcomes for different combinations of effect sizes and assuming a ICC of 0.05, a CAC of 0.8, and a type I error rate of 0.05. Power for non-linear models calculated using a normal approximation.

| **Outcome** | **Assumed model** | **Assumed baseline** | **Effect size** | | **Obs. per cluster-period** | **Power** |
| --- | --- | --- | --- | --- | --- | --- |
| *Analysis 1* | | | | | | |
| Water and Food safety and hygiene behaviour | Binomial-logistic | 50% | +5pp | | 27 mother-child pairs  4 opportunities per pair | 57% |
|  |  |  | +10pp | |  | >99% |
|  |  |  | +20pp | |  | >99% |
| Food and water contamination | Poisson | 10 cfu/g | -1 | | 10 samples per cluster | >99% |
|  |  |  | -2 | |  | >99% |
| Diarrhoea | Binomial-logistic | 13% | -2pp | | 27 mother-child pairs | 26% |
|  |  |  | -3pp | |  | 52% |
|  |  |  | -5pp | |  | 94% |
|  |  |  | -7pp | |  | >99% |
| *Analysis 2: Subgroup analyses* | | | | | | |
| **Outcome** | **Assumed model** | **Assumed baseline** | **Main effect** | **Interaction effect** | **Obs. per cluster-period** | **Power** |
| Water and Food safety and hygiene behaviour | Binomial-logistic | 50% | 0pp | +5pp | 27 mother-child pairs  4 opportunities per pair | 18% |
|  |  |  | 0pp | +10pp |  | 57% |
|  |  |  | +5pp | +5pp |  | 19% |
|  |  |  | +5pp | +10pp |  | 59% |
| Food and water contamination | Poisson | 10 cfu/g | 0 | -1 | 10 samples per cluster | 25% |
|  |  |  | 0 | -2 |  | 78% |
|  |  |  | -2 | -1 |  | 37% |
|  |  |  | -2 | -2 |  | 93% |
| Diarrhoea | Binomial-logistic | 13% | 0pp | -2pp | 27 mother-child pairs | 10% |
|  |  |  | 0pp | -5pp |  | 44% |
|  |  |  | -2pp | -2pp |  | 11% |
|  |  |  | -2pp | -5pp |  | 55% |
|  |  |  | -5pp | -2pp |  | 15% |

**Table S3** Power for the three main outcomes for different combinations of effect sizes and assuming a ICC of 0.05, a CAC of 0.5, and a type I error rate of 0.05. Power for non-linear models calculated using a normal approximation.

| **Outcome** | **Assumed model** | **Assumed baseline** | **Effect size** | | **Obs. per cluster-period** | **Power** |
| --- | --- | --- | --- | --- | --- | --- |
| *Analysis 1* | | | | | | |
| Water and Food safety and hygiene behaviour | Binomial-logistic | 50% | +5pp | | 27 mother-child pairs  4 opportunities per pair | 50% |
|  |  |  | +10pp | |  | 98% |
|  |  |  | +20pp | |  | >99% |
| Food and water contamination | Poisson | 10 cfu/g | -1 | | 10 samples per cluster | >99% |
|  |  |  | -2 | |  | >99% |
| Diarrhoea | Binomial-logistic | 13% | -2pp | | 27 mother-child pairs | 23% |
|  |  |  | -3pp | |  | 46% |
|  |  |  | -5pp | |  | 89% |
|  |  |  | -7pp | |  | >99% |
| *Analysis 2: Subgroup analyses* | | | | | | |
| **Outcome** | **Assumed model** | **Assumed baseline** | **Main effect** | **Interaction effect** | **Obs. per cluster-period** | **Power** |
| Water and Food safety and hygiene behaviour | Binomial-logistic | 50% | 0pp | +5pp | 27 mother-child pairs  4 opportunities per pair | 16% |
|  |  |  | 0pp | +10pp |  | 51% |
|  |  |  | +5pp | +5pp |  | 17% |
|  |  |  | +5pp | +10pp |  | 53% |
| Food and water contamination | Poisson | 10 cfu/g | 0 | -1 | 10 samples per cluster | 24% |
|  |  |  | 0 | -2 |  | 76% |
|  |  |  | -2 | -1 |  | 36% |
|  |  |  | -2 | -2 |  | 92% |
| Diarrhoea | Binomial-logistic | 13% | 0pp | -2pp | 27 mother-child pairs | 9% |
|  |  |  | 0pp | -5pp |  | 39% |
|  |  |  | -2pp | -2pp |  | 10% |
|  |  |  | -2pp | -5pp |  | 48% |
|  |  |  | -5pp | -2pp |  | 14% |
